# A Novel Computer-Assisted Drug-Induced Liver Injury Causality Assessment Tool (DILI-CAT)

**DOI:** 10.1101/2021.03.05.21252982

**Authors:** Hans L. Tillmann, Ayako Suzuki, Michael Merz, Richard Hermann, Don C. Rockey

**Author notes:** **Corresponding author:** Hans L. Tillmann, MD, Division of Gastroenterology, Hepatology &, Nutrition, East Carolina University, 600 Moye Blvd., Greenville, NC 27834, USA.

## Abstract

**Objective:** We hypothesized that a drug-characteristic DILI-phenotype could be defined and be used to develop a computer-assisted DILI causality assessment-tool (DILI-CAT).

**Design:** A drug-specific DILI-phenotype was developed for amoxicillin-clavulanate (AMX/CLA), cefazolin, cyproterone, and polygonum multiflorum using data from published case series, and subsequently a DILI-CAT Score (DILI-CAT-S) was created for each drug. The phenotype was made up of the following three parameters: (1) latency, R-value, and (3) AST/ALT ratio (also de Ritis ratio). A point allocation system was developed with points allocated depending on the degree of deviation from the core of published data for the three phenotypic parameters.

**Results:** The four drugs had a significantly different phenotype based on the three parameters utilized. For example, the median cyproterone latency was 150 days versus less than 43 days for the other three drugs (median: 26 for AMX/CLA, 20 for cefazolin, and 20 days for polygonum multiflorum; p<0·001). The R-value for the four drugs was also significantly different (median: cyproterone [12.4] and polygonum multiflorum [10.9]) from AMX/CLA (1.44) and cefazolin (1.57; p<0.001). The resulting DILI-CAT-S effectively separated cyproterone and polygonum multiflorum from AMX/CLA and cefazolin, respectively (p<0·001). Notably, because of overlap in phenotype AMX/CLA and cefazolin could not be differentiated by DILI-CAT-S.

**Conclusion:** DILI-CAT is a data-driven, diagnostic tool built to define drug-specific phenotypes. Data presented here provide proof of principle that a drug-specific, data-driven causality assessment tool can be developed for different drugs and raise the possibility that such a process could improve and standardize causality assessment methods.

**Funding:** DCR was supported by the NIH, grant P30 DK 123704

## Introduction

Drug-induced liver injury (DILI) is an important cause of acute liver injury and liver-related morbidity and mortality.^1-4^ DILI is also a major concern in drug development and post-marketing surveillance, as evidenced by hepatotoxicity being a leading cause for market withdrawal of licensed drugs.^5^ Moreover, DILI diagnosis is extremely challenging, as liver biochemistry abnormalities may be caused by a variety of different causes of liver injury.^6-8^

Unlike diseases such as viral hepatitis, where diagnostic testing may confirm or exclude the diagnosis with high sensitivity and specificity, DILI is a diagnosis based on specific clinical features associated with a particular drug and requires exclusion of other causes of liver diseases. A variety of causality assessment methods (CAMs) have been developed and often use point-scoring systems (ie, Roussel Uclaf Causality Assessment Method [RUCAM], “clinical diagnostic scale” [CDS]).^9-11^ While these scoring systems have attempted to quantify the causality likelihood for DILI, a structured expert opinion process (SEOP), such as described by the Drug Induced Liver Injury Network (DILIN), has been shown to be superior to RUCAM.^12^ We have previously shown that different drugs have different clinical DILI characteristics or phenotypes.^13^ We believe that one of the reasons why SEOP is superior to RUCAM is that experts recognize specific clinical phenotypes (ie, its “signature” or typical characteristics) for different drugs that cause DILI. Unfortunately, a major limitation of the expert opinion approach is that it is not widely available in clinical practice and is thus not generalizable. It is likely that one reason why the expert opinion approach is successful is that experts have great experience with DILI and are often familiar with drug-specific DILI phenotypes.

Here, we hypothesized that not only are there clinical DILI features that are typical for each drug and make up a typical phenotype, but also that such drug-specific DILI phenotypes could be used to develop a novel DILI causality assessment tool (DILI-CAT), incorporating data-driven drug-specific DILI phenotypes. We aimed to create a quantitative data-driven algorithm (DILI-CAT) to define drug-specific DILI phenotypes using characteristic DILI features. These include (1) latency, (2) R-value (the ratio of alanine aminotransferase [ALT] to the upper limit of normal for [ALT] / alkaline phosphatase [ALP] to the upper limit of normal for ALP] and (3) the aspartate aminotransferase [AST] / ALT ratio [de Ritis ratio] derived from available literature that included patient-level data. We also sought to develop drug-specific scoring systems to quantify the resemblance of acute liver injury events to the defined DILI phenotypes.

## Methods

We performed a literature search to identify published case-series studies reporting clinical features in patients with DILI caused by single specific drugs. Although most studies that were identified failed to report patient-level data necessary to develop a robust DILI phenotype, we identified four case series that fulfilled the requirement of having detailed patient-level data for latency, R-value, and AST/ALT ratio (de Ritis) at onset. These included one study for each of the following four drugs: Cyproterone (n=22),^14^ amoxicillin-clavulanate (n=35),^15^ cefazolin (n=19),^16^ and polygonum multiflorum (n=18).^17^

### Design

We considered the drug-specific DILI phenotype to be made up primarily of its latency (in days), absolute R-value, and AST/ALT ratio. A quantitative “drug-specific” scoring system was then developed that allocates points based on these three specific DILI features. For DILI-CAT scoring, a drug-specific DILI-CAT Score (DILI-CAT-S) would be developed based on the distribution of the respective latencies, R-values, and AST/ALT ratios in identified case series. A separate DILI-CAT-S was developed for each drug. Furthermore, depending on the difference in phenotype being more marked in latency, R-value, or AST/ALT ratio, the respective DILI-CAT-S was weighted in the respective category, resulting in latency-weighted DILI-CAT-S, R-value□weighted DILI-CAT-S, or AST/ALT ratio□weighted DILI-CAT-S. The category of the three (latency, R-value, and AST/ALT ratio) in which the greatest differences between two drugs in non-parametric comparison were detected received two-fold greater weight compared to the remaining two categories.

We hypothesized that the closer a drug’s DILI features (ie, latency, R-value, and AST/ALT ratio) are to the published clinical characteristics, the more likely the case is to be a bona fide DILI case. In other words, the closer a case’s values are to the interquartile range (IQR) of values in published DILI cases for that drug, the more likely that injury is related to the drug in question.

In the model, points were allocated based on the closeness of the parameter in question (latency, R-value, AST/ALT ratio) for each specific drug to the IQR, or 50% core interval (Figure 1, Supplemental Figure 1a & 1b) as derived from known cases (patient/case level data).^14-17^ Proportionally fewer points were allocated when values for the parameter in question fell outside the IQR. By definition, there was variability in points allocated for each drug at different latency, R-value, and AST/ALT ratio because allocated points were dependent on the respective 50% core interval (IQR) and percentiles for each drug (Table 1 and Supplemental Table 1). Any parameter falling within the core interval was allocated 20 points. Values outside the 50% core interval (IQR) were given fewer points (Table 1). Deductions were given for values outside of the range of the values for respective drug’s phenotype range (Figure 1, Table 2, Supplemental Table 1). Additional deductions were also given when values were far outside the IQR; these were defined as “outliers” (Supplemental Material Appendix 1).

**Figure 1.**
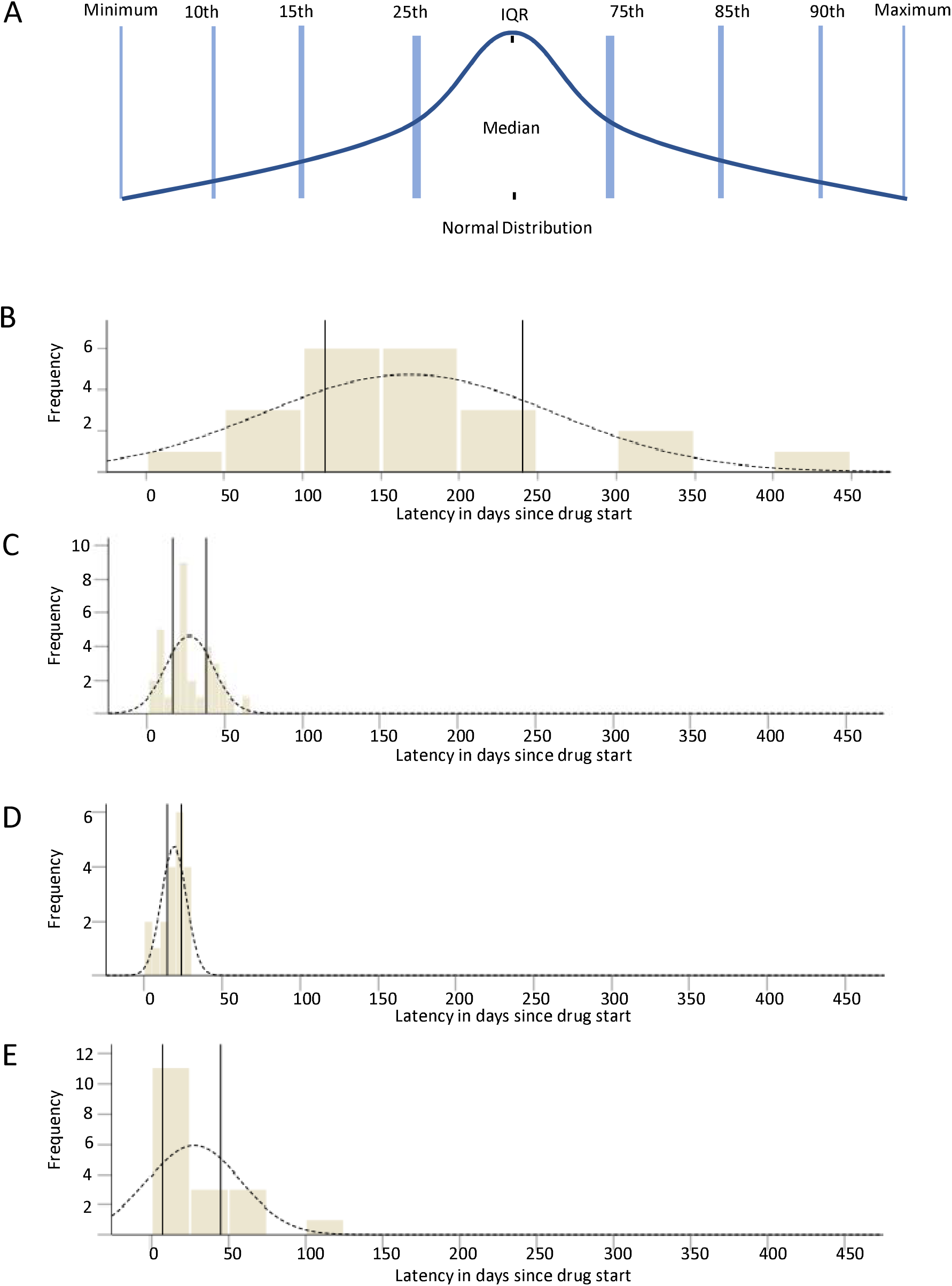
Frequency distribution of latency of cases compared to a normal distribution. Normal distribution (A) is shown compared to distribution of non-normal distribution of latency among DILI cases due to cyproterone (B), AMX/CLA (C), cefazoline (D), and polygonum multiflorum (E). In panels B to E, frequency of cases is given on the Y axis and latency in days from drug start in the X axis; the vertical lines in panels B-E represent the interquartile range or 25^th^ and 75^th^ percentile. AMX/CLA, amoxicillin/clavulanate; DILI, drug-induced liver injury.

**Table 1:** **Relative point allocation according to value relative to the distribution of values within the respective case series**

|  | **Percentage of points to be allocated** | **Point allocation** |
| --- | --- | --- |
| **IQR low-IQR high** | 100% | 20 |
| **25<sup>th</sup> to 15<sup>th</sup> percentile** | 50% | 10 |
| **75<sup>th</sup> to 85<sup>th</sup> percentile** |  |  |
| **15<sup>th</sup> to 10<sup>th</sup> percentile** | 25% | 5 |
| **85<sup>th</sup> to 90<sup>th</sup> percentile** |  |  |
| **10<sup>th</sup> percentile to minimum of range** | 0% | 0 |
| **90<sup>th</sup> percentile to maximum of range** |  |  |
| **Below minimum of range** | -25% | -5 |
| **Above maximum of range** |  |  |
| **Outlier\*** | -25% | -5 |

\* For cases with values that are outside the range and are outliers, -50% is used. IQR, interquartile range.

**Table 2.**
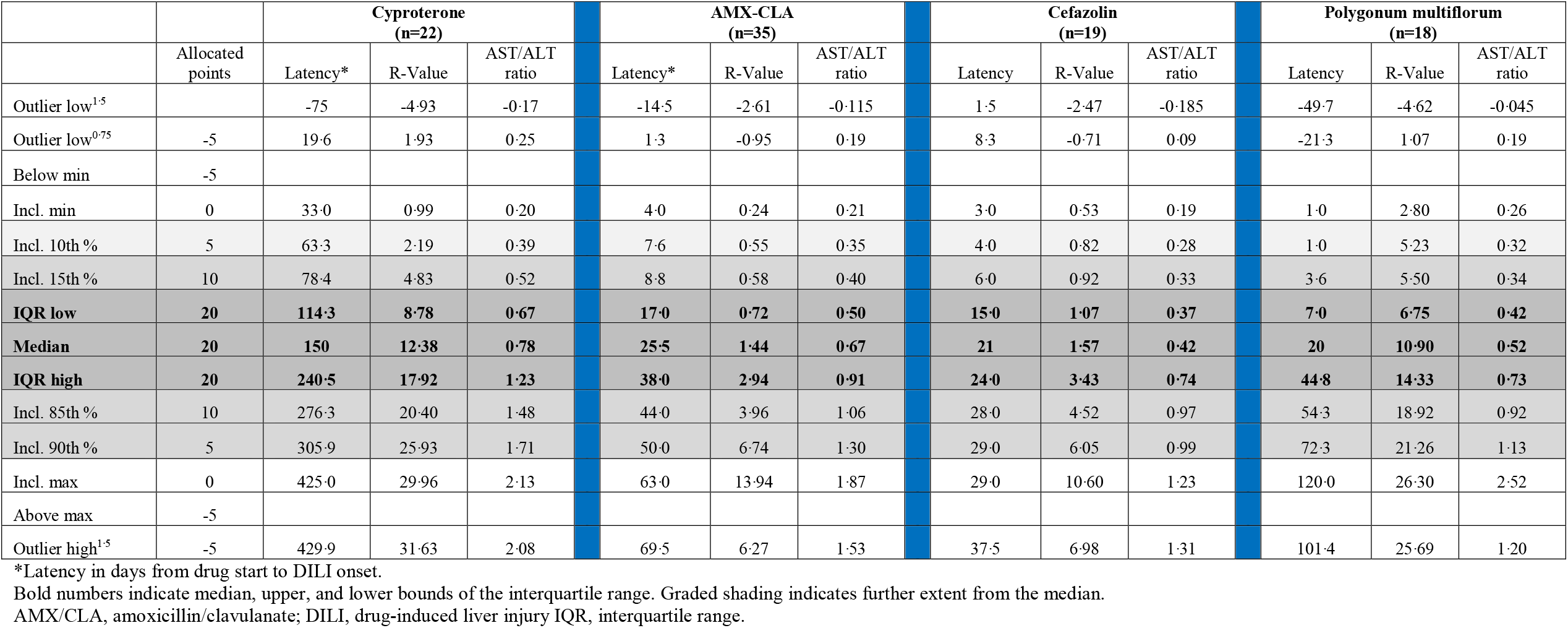
**Drug phenotypes described by Latency, R-Value & AST/ALT ratio with interquartile range and percentiles**

The strategy that was ultimately utilized to generate a scoring system required several assumptions. First, we postulated that each of the four drugs chosen (or any other drug, for that matter) would exhibit differences in one or more of the three clinical categories (latency, R-value, AST/ALT ratio) compared to at least some other drugs. We also postulated that the clinical categories that exhibited the greatest differences would be the most valuable in differentiating clinical phenotypes between drugs (ie, latency for one drug would be much longer than the others, and therefore was highly valuable in defining the clinical phenotype). For the specific category with the greatest discriminating potential (latency, R-value, AST/ALT ratio), that category’s value was doubled. For example, if for a specific drug, latency exhibits the greatest statistical difference compared to R-value and AST/ALT ratio differences, then latency points were doubled.

### Statistics

Each drug’s phenotype was informed by the IQR, percentiles, maximum and minimum values, and definition of outlier values for each of the three clinical variables (latency, R-value, AST/ALT ratio).

A non-parametric Mann-Whitney rank test was used to compare drug phenotypes (for each of the three clinical variables latency, R-value, AST/ALT ratio) to each other. Differences in variables were defined statistically (Table 3). The smaller the Mann-Whitney value, the greater the difference, and a Mann-Whitney U value of “0” reflects complete separation of parameters between groups (ie, the Mann-Whitney number comparing latency for cyproterone and cefazolin was zero, reflecting that all latencies for cyproterone were longer than any cefazolin latencies).

**Table 3.** **Differences in Phenotypes Categories (Latency, R-Value & AST/ALT ratio) between drugs**

|  | Cyproterone (n=22) |  |  |  | AMX-CLA (n=35) |  |  |  | Cefazolin (n=19) |  |  |  | Polygonum multiflorum (n=18) |  |  |
| --- | --- | --- | --- | --- | --- | --- | --- | --- | --- | --- | --- | --- | --- | --- | --- |
|  | Latency* | R-Value | AST/ALT ratio |  | Latency* | R-Value | AST/ALT ratio |  | Latency* | R-Value | AST/ALT ratio |  | Latency* | R-Value | AST/ALT ratio |
| **Cyproterone** |  |  |  |  |  |  |  |  |  |  |  |  |  |  |  |
| Mann-Whitney U | NA | NA | NA |  | **12·5** | 65·0 | 289·0 |  | **0** | 34 | 100 |  | **12** | 164 | 101 |
| p-value | NA | NA | NA |  | **<0·000** | 0·000 | 0·116 |  | **<0·000** | 0·000 | 0·004 |  | **<0·000** | 0·355 | 0·008 |
| **AMX-CLA** |  |  |  |  |  |  |  |  |  |  |  |  |  |  |  |
| Mann-Whitney U | **12·5** | 65 | 289 |  | NA | NA | NA |  | 227·5 | 276 | **218** |  | 262·000 | **41** | 232 |
| p-value | **<0·000** | <0·000 | 0·116 |  | NA | NA | NA |  | 0·057 | 0·306 | **0·038** |  | 0·319 | **<0·000** | 0·119 |
| **Cefazolin** |  |  |  |  |  |  |  |  |  |  |  |  |  |  |  |
| Mann-Whitney U | **0** | 34 | 100 |  | 227 | 276 | **218** |  | NA | NA | NA |  | 171 | **17** | 146 |
| p-value | **<0·000** | <0·000 | 0·004 |  | 0·057 | 0·306 | **0·038** |  | NA | NA | NA |  | 1·000 | **<0·000** | 0·447 |
| **Polygonum multiflorum** |  |  |  |  |  |  |  |  |  |  |  |  |  |  |  |
| Mann-Whitney U | **12** | 164 | 101 |  | 262 | **41** | 232 |  | 171 | **17** | 146 |  | NA | NA | NA |
| p-value | **<0·000** | 0·355 | 0·008 |  | 0·319 | **<0·000** | 0·119 |  | 1·000 | **<0·000** | 0·447 |  | NA | NA | NA |

\*Latency in days from drug start to DILI onset.
Bold numbers in shaded areas represent the most significant differences.
AMX/CLA, amoxicillin/clavulanate; DILI, drug-induced liver injury; NA, not applicable.

For each drug, a drug-specific DILI-CAT-S was developed using a scoring algorithm. To compare drug-specific DILI-CAT-S’ performance, each drug was evaluated using its respective DILI-CAT-S against the three other drugs, where the significance of difference was assessed using the Mantel-Haenszel test for trend considering five-point incremental scores as ordinal categories. Data handling was done using Microsoft^®^ Excel^®^, and IBM^®^ SPSS^®^ version 25 was used for statistical analysis.

## Results

The typical latency for cyproterone was considerably longer (median 150 days, IQR 114-240 days) than that for the other three drugs (which ranged from median 20 to median 26 days; IQR 16,·25-24 days to 7-44 days; Table 2, Figure 1, Supplemental Figure 1). The R-values also varied but at the same time were similar for some of the drugs, with median values of 10·9 (IQR 7·6-13·5) and 12·4 (IQR 9·83-17·84) for polygonum multiflorum and cyproterone, respectively, and 1·4 (IQR 0·74-2·92) and 1·6 (IQR 1·07-2·87) for AMX-CLA and cefazolin, respectively (Table 2). The AST/ALT ratios largely overlapped among all these four drugs (from 0·52 to 0·78).

### Phenotypic differences among drugs

The DILI-CAT-S utilizes weighting of individual phenotypic features (latency, R-value, and AST/ALT ratio, as described in the Methods).

Cyproterone showed the greatest difference in latency compared to the other three drugs (p<0·001, Table 3). While polygonum multiflorum differed from cyproterone most strongly in terms of latency (p<0·001, Table 3) and to lesser extent in AST/ALT ratio (p=0·008, Table 3), polygonum multiflorum differed from AMX-CLA and cefazolin significantly only in R-value (p<0·001, Table 3). AMX-CLA and cefazolin differed only mildly in the AST/ALT ratio from each other (p=0·038, Table 3). Based on the respective greatest difference, as defined by lowest U-value (Table 3), the following weighting was applied:

- For cyproterone a latency-weighted (thus latency valued double) DILI-CAT was applied for comparison against all other three drugs.
- For polygonum multiflorum, a latency-weighted DILI-CAT was applied for comparison against cyproterone, but an R-value□weighted DILI-CAT was applied for comparison against both AMX-CLA and cefazolin.
- For both AMX-CLA and cefazolin, a latency-weighted DILI-CAT was applied against cyproterone, an R-value□weighted DILI-CAT against MP, and, finally, AMX-CLA and cefazoline were compared using an AST/ALT ratio□weighted DILI-CAT.

### Cyproterone-derived DILI-CAT

In order to create a DILI-CAT score, as outlined in the Methods, points were allocated based on latency, R-value, and AST/ALT ratios. As predetermined, a latency-weighted DILI-CAT would be applied that shows significant difference against all three other drugs (Table 4, Supplemental Figure 2a).

**Table 4:** **Drugs’ phenotype-derived DILI-CAT scores show potential for liver injury event separation dependent on drug and applied drug-characteristic DILI-CAT derived from phenotype as outlined in Table 1**

**Cyproterone vs. other drugs using DILI-CAT-S<sub>Cyproterone</sub> median points**
|  | Categories |  |  | DILI-CAT |  |  |
| --- | --- | --- | --- | --- | --- | --- |
|  | Latency | R-Value | de Ritis | Latency weighted | R-Value weighted | de Ritis weighted |
| **Cyproterone (n=22)** | **20** | **20** | **20** | **47.5** | **55** | **50** |
| AMX-CLA (n=35) | -5 | -5 | 10 | 5 | 0 | 20 |
| Cefazolin (n=19) | -5 | -5 | 10 | 5 | 5 | 20 |
| PM (n=18) | -7.5 | 10 | 10 | 15 | 32.5 | 30 |
| P-values for Cyproterone versus | Mantel-Haenszel test for trend |  |  |  |  |  |
| AMX-CLA (n=35) | **<0.001** | **<0.001** | 0.785 | **<0.001** | **<0.001** | **<0.001** |
| Cefazolin (n=19) | **<0.001** | **<0.001** | **0.595** | **<0.001** | **<0.001** | **<0.001** |
| PM (n=18) | **<0.001** | 0.984 | 0.352 | **<0.001** | **0.004** | **0.001** |

**AMX-CLA vs. other drugs using DILI-CAT-S<sub>AMX/CLA</sub> median points**
|  | Categories |  |  | DILI-CAT |  |  |
| --- | --- | --- | --- | --- | --- | --- |
|  | Latency | R-Value | de Ritis | Latency weighted | R-Value weighted | de Ritis weighted |
| Cyproterone (n=22) | -10 | -5 | 20 | -7.5 | -2.5 | 20 |
| **AMX-CLA (n=35)** | **20** | **20** | **20** | **55** | **60** | **60** |
| Cefazolin (n=19) | 20 | 20 | 10 | 70 | 60 | 60 |
| PM (n=18) | 2.5 | -5 | 15 | 25 | 10 | 27.5 |
| P-values for AMX-CLA versus | Mantel-Haenszel test for trend |  |  |  |  |  |
| Cyproterone (n=22) | **<0.001** | **<0.001** | 0.785 | **<0.001** | <0.001 | <0.001 |
| Cefazolin (n=19) | 0.357 | 0.379 | 0.356 | 0.453 | 0.496 | 0.972 |
| PM (n=18) | **0.003** | **<0.001** | 0.989 | <0.001 | **<0.001** | <0.001 |

**Cefazolin vs. other drugs using DILI-CAT-S<sub>Cefazolin</sub>, median points**
|  | Categories |  |  | DILI-CAT |  |  |
| --- | --- | --- | --- | --- | --- | --- |
|  | Latency | R-Value | de Ritis | Latency weighted | R-Value weighted | de Ritis weighted |
| Cyproterone (n=22) | -10 | -10 | 10 | -12.5 | -10 | 0 |
| AMX-CLA (n=35) | 10 | 10 | 20 | 30 | 40 | 40 |
| **Cefazolin (n=19)** | **20** | **20** | **20** | **60** | **60** | **60** |
| PM (n=18) | 0 | -7.5 | 20 | 17.5 | 10 | **25** |
| P-values for Cefazolin versus | Mantel-Haenszel test for trend |  |  |  |  |  |
| Cyproterone (n=22) | **<0.001** | **<0.001** | **0.045** | **<0.001** | **<0.001** | **<0.001** |
| AMX-CLA (n=35) | **0.005** | 0.082 | 0.244 | **0.002** | **0.009** | **0.008** |
| PM (n=18) | **0.002** | **<0.001** | 0.990 | **<0.001** | **<0.001** | **<0.001** |

**Polygonum Multiflorum vs. other drugs using DILI-CAT-S<sub>Polygonum Multiflorum</sub>, median points**
|  | Categories |  |  | DILI-CAT |  |  |
| --- | --- | --- | --- | --- | --- | --- |
|  | Latency | R-Value | de Ritis | Latency weighted | R-value weighted | de Ritis weighted |
| Cyproterone (n=22) | -10 | 15 | 5 | 7.5 | 32.5 | 25 |
| AMX-CLA (n=35) | 20 | -5 | 10 | 45 | 20 | 40 |
| Cefazolin (n=19) | 20 | -5 | 10 | 45 | 20 | 35 |
| **PM (n=18)** | **20** | **20** | **20** | **57.5** | **60** | **57.5** |
| P-values for PM versus | Mantel-Haenszel test for trend |  |  |  |  |  |
| Cyproterone (n=22) | **<0.001** | 0.282 | **0.033** | **<0.001** | **0.001** | **<0.001** |
| AMX-CLA (n=35) | 0.031 | <0.001 | 0.169 | 0.011 | **<0.001** | 0.002 |
| Cefazolin (n=19) | 0.084 | <0.001 | 0.07 | 0.016 | **<0.001** | 0.001 |

AMX/CLA, amoxicillin/clavulanate; DILI-CAT, drug-induced liver injury causality assessment tool; DILI-CAT-S, drug-induced liver injury causality assessment tool score; PM: polygonum multiflorum.

### AMX/CLA-derived DILI-CAT

AMX/CLA had a more complex clinical pattern; the most significant differences found in the three clinical categories depended on the comparator drug. For example, latency was most distinct from cyproterone (Table 3), R-value most different from polygonum multiflorum (Table 3), and AST/ALT ratio most distinct from cefazolin (Table 3). Latency weighting provided the greatest discrimination between AMX-CLA and cyproterone, while an R-value□weighted DILI-CAT provided the greatest discrimination between AMX-CLA and polygonum multiflorum (Table 4). In contrast, AST/ALT ratio weighting did not provide added discriminatory value (Table 4, Supplemental Figure 2b).

### Cefazolin-derived DILI-CAT performance

Cefazolin was similar to AMX/CLA in all three clinical categories. It had the greatest difference in latency compared to cyproterone and was most distinct in R-value compared to polygonum multiflorum. Its AST/ALT ratio was most distinct from AMX/CLA (though minimally different [p = 0·038; Table 3]).

While the AMX/CLA-derived DILI CAT did not separate AMX/CLA from cefazolin, a cefazolin-derived DILI-CAT was able to better separate cefazolin from AMX/CLA (p=0·008; Table 4, Supplemental Figure 2c). This is likely because the cefazolin phenotype shows less variation in latency, R-value and AST/ALT ratio compared to the AMX/CLA phenotype, where more AMX/CLA cases overlap with cefazolin’s phenotype than vice versa (Table 2).

### Polygonum multiflorum □ derived DILI-CAT performance

Polygonum multiflorum was most different from cyproterone in the latency category, while the latency of polygonum multiflorum and either AMX/CLA or cefazolin was similar (Table 3). Polygonum multiflorum’s R-value was distinct from AMX/CLA and cefazolin (Table 3). Therefore, a latency-weighted polygonum multiflorum□derived DILI-CAT shows the greatest separation from cyproterone (Table 4), while an R-value□weighted DILI-CAT shows the greatest separation against AMX/CLA as well as cefazolin (Table 4, Supplemental Figure 2d).

## Discussion

Here, we have developed a data-driven approach (DILI-CAT) that can be used to create a drug-specific DILI phenotype and enhance DILI causality assessment. We demonstrate that drug phenotypes differ significantly and that our algorithmic approach allows for differentiation of DILI caused by different drugs.

RUCAM, the commonly used causality assessment method (tool) developed almost three decades ago,^9^ is often considered the most reliable approach to DILI causality assessment when an expert opinion assessment is not available.^12^ However, neither RUCAM nor any of the other currently available causality assessment tools uses a drug-specific approach.^18^ An SEOP is considered superior to RUCAM,^1,2^ which is likely due to the fact that experts probably consider a drug’s phenotype even when there is no mechanism to quantitate the influence of a drug-specific phenotype. Implicit in the findings presented here is that allowing in a formal process for inclusion of a drug phenotype enhances the DILI adjudication process by including phenotypic characteristics of drug-specific DILI. We speculate that this will be helpful to experts and, perhaps to an even greater degree, to nonexperts.^19^

An algorithmic data-driven and drug-specific diagnostic tool such as DILI-CAT has a number of advantages. DILI-CAT’s data-driven approach is objective. It can be optimized via weighting of specific variables, which will allow for better discrimination between different drugs. The intrinsic propensity for hepatotoxicity of a drug (ie, the likelihood or probability that a specific drug would cause liver injury) could be included in the mathematical algorithm (Supplement Appendix 2, 3, and 4; Supplement Table 2), and could be readily derived from the published literature.^20,21^ Scoring for competing causes in DILI-CATs could also be included, allowing for grading of individual drugs along a causality scale (Supplemental Appendix 2). Further, the flexible format of DILI-CAT allows it to be programmed for use by any drug, as long as the DILI phenotype of a drug can be characterized (eg, with a sufficient number of known DILI cases to estimate percentiles of the drug-specific features). Finally, the approach should be considered to be a “living” process, meaning that additional cases could be added as more published cases become available so as to create a more robust DILI signature.

We recognize limitations of the current version of DILI-CAT. One limitation is that some drugs will have overlapping phenotypes, such as was the case with cefazolin and AMX/CLA; in this situation, DILI-CAT-s will be unable to provide a clear distinction between a drug in question and other competing drugs. Another limitation is that DILI-CAT depends on having available cases with which to develop specific drug signatures. In this regard, we speculate that the number of cases needed to develop a robust signature will depend on the consistency of the drug’s phenotype.^19^ The more variable the phenotype, the more cases that are likely to be required to generate a precise picture of a drug’s signature.

In the future, we envision a staggered approach to DILI causality assessment. First, the likelihood of DILI could be assessed using an algorithmic methodology such as that presented here, and secondly laboratory testing could be used for confirmation. While lymphocyte transformation test (LTT) is recommended in the Japanese DDW-J scoring for DILI,^22^ it is unclear whether this assay is reproducible enough to be used.^23^ A novel promising approach is based on assay of blood derived monocytes that are transformed into hepatocyte like cells.^24^ In a number of studies, this test has shown promise as a confirmatory assay.^25-27^

In summary, we have presented an objective and data-driven drug-specific tool (DILI-CAT) that represents a novel and substantial step forward in DILI causality assessment. This approach is likely to be extremely useful for clinicians who are not experts in DILI causality assessment, and it also has the potential to improve expert adjudication of DILI.

## Data Availability

Analysis only includes publicly available data.

https://doi.org/10.1111/liv.12899

https://doi.org/10.1016/s0016-5085(99)70404-x

https://doi.org/10.1016/j.cgh.2014.11.036

https://doi.org/10.1016/j.ctim.2013.12.008

## Contributors

All authors - study concept and design; acquisition of data; interpretation of data; drafting of the manuscript; critical revision of the manuscript for important intellectual content.

Hans L. Tillmann and Richard Hermann have verified the underlying data.

Hans L. Tillmann - data analysis

Richard Hermann - project administration, supervision

Don C. Rockey - literature review figures, data analysis

## Declaration of interests

Hans L. Tillmann is a stockholder of Abbott, AbbVie, and Gilead outside the submitted work. He reports that his wife is a full-time employee of AbbVie.

Ayaka Suzuki has nothing to disclose.

Michael Merz has nothing to disclose.

Richard Hermann is a full-time employee and stockholder at AstraZeneca.

Don C. Rockey has nothing to disclose.

## Acknowledgments

Dr. Huiman Barnhart was instrumental in developing earlier versions of this approach. Editorial assistance, funded by AstraZeneca, was provided by Peloton Advantage, LLC, an OPEN Health company. Additional statistical support was provided by Daniel Chima, an employee of AstraZeneca.

## Data Statement

The data used here from published case series with data derived from the published articles, as the patient level data were derived from published cares series. Data will also be available with publication by outreach to the corresponding author and will be shared for analyses to replicate findings, after approval of a proposal and with a signed data access agreement, at a minimum.

## Patient and Public Involvement statement

Due to the nature of this research that did not involve direct patient contact, not patient and public involvement was used for this study.

