## Supplementary material for "A Novel Computer-Assisted Drug-Induced Liver Injury Causality Assessment Tool (DILI-CAT)": Supplemendal

**Appendix 1**

**Outlier definition:**
There can be outliers on either side of the Interquartile Range (IQR). "High-end outliers," or values above the IQR, were defined as values that were equal to or greater than the upper end of the IQR (equal to the top end of the 3rd quartile) plus 1·5 times the IQR ($IQR_{high}$+IQR*1·5).

Using a similar definition, the "low-end outlier" would mathematically allow even negative values that are impossible for the phenotype features (latency for drug-induced liver injury onset cannot occur before a drug is started, R-value [ratio of alanine aminotransferase (ALT) in upper limit of normal (ULN) / alkaline phosphatase in ULN, where a ratio cannot be negative], and the aspartate aminotransferase [AST] /ALT ratio [which likewise cannot be negative]). In order to avoid or minimise "low-end outlier definitions" from falling into a negative value, "low-end outliers were defined as "low-end IQR minus IQR* 0·75" ($IQR_{low}$-IQR*0·75). Therefore, low-end outlier would mostly be avoided.

**Appendix 2**

**Hepatotoxicity potential (0-20 points)**

Drugs have different propensities for causing drug-induced liver injury (DILI). *LiverTox* (available at Pubmed.ncbi.nlm.nih.gov) provides an estimate of the propensity that a certain drug will cause DILI and, thus, we used this scoring system. This system can be used for the following point allocations:

Drugs with ≥ 50 cases reported in the literature = 20 points
Drugs with ≥ 12 but < 50 cases reported in the literature = 15 points
Drugs with ≥ 4 but <12 cases reported in the literature = 10 points
Drugs with 1 to 3 cases reported in the literature = 10 points
Drugs with no cases reported in the literature = 0 points

### Appendix 3

**Point allocation for competing causes and the lack thereof (-25 to 25 points)**

For this project we had a few "a priori" considerations:

1. If all alternative causes were truly excluded, the score should, at minimum, result in a score of possible, even when the respective drug was not known to be hepatotoxic, and therefore no phenotype of the drug is known.

2. Even if an alternative cause is identified but a case's injury pattern completely fits the expected phenotype, the resulting score should not exclude drug-induced liver injury (DILI) completely.

Thus, if a definite alternative cause of abnormal liver tests was identified, 25 points were to be deducted. If no alternative cause was positively identified, completeness of exclusion of alternative causes is important, since DILI is a diagnosis of exclusion. We arbitrarily allocated 25 points if all relevant competing causes were excluded under the premise that a case with suspected DILI to a drug with no previous report of hepatotoxicity should be adjudicated as possible DILI. Depending on the injury pattern, hepatocellular vs. cholestatic, different diseases needed to be excluded. In the case of a mixed injury pattern, both causes of hepatocellular and cholestatic injury needed to be excluded. If not all compatible diseases were excluded, various points were deducted from the 25 points depending on a weighted importance of a competing cause to be excluded.

Diseases differ in their importance to be excluded. Thus, a relative exclusionary importance of various causes of liver disease was developed (Supplemental Table 2). The resulting point deduction from 25 potential points was derived from the respective score for the exclusion of the formular 25 minus 25 points multiplied by the relative importance of missing excluded diagnosis from 25 points. If the sum of relative importance of missing values excluded competing diagnosis was between 1 and <2, a relative importance value of 1 was used. If relative importance of missing values was ≥2 a relative value of 1·4 was used (see examples for details).

<u>Examples:</u>

- If the sum of relative importance of missing values was 0·3, a total of 7·5 points were deducted from 25 points, resulting in a net value of 17.5 points to be added to the score.
- If the sum of relative importance of missing values was 0·7, a total of 17·5 points were deducted from 25 points, resulting in a net value of 7·5 points to be added to the score.
- If the sum of relative importance of missing values was between 1 and <2, a relative importance value of 1 was used, resulting in 25 points to be deducted from 25 points, resulting in a net value of no point to be added to the score.
- If the sum of relative importance of missing values was ≥2, 35 points were to be deducted from the 25 points, resulting in a net value of 10 points to be subtracted from the total score.

**Appendix 4**

The summation of points in the three categories (DILI-CAT phenotype, hepatotoxicity, and competing causes) theoretical ranges from -65 [drug with no history of hepatotoxicity (0 points), completely not fitting the signature (-40 points) and positive alternative cause (-25 points)] to 125 [drug known to be frequently hepatotoxic (20 points), complete fit with DILI-CAT phenotype (80 points), and all alternative causes excluded (25 points)].

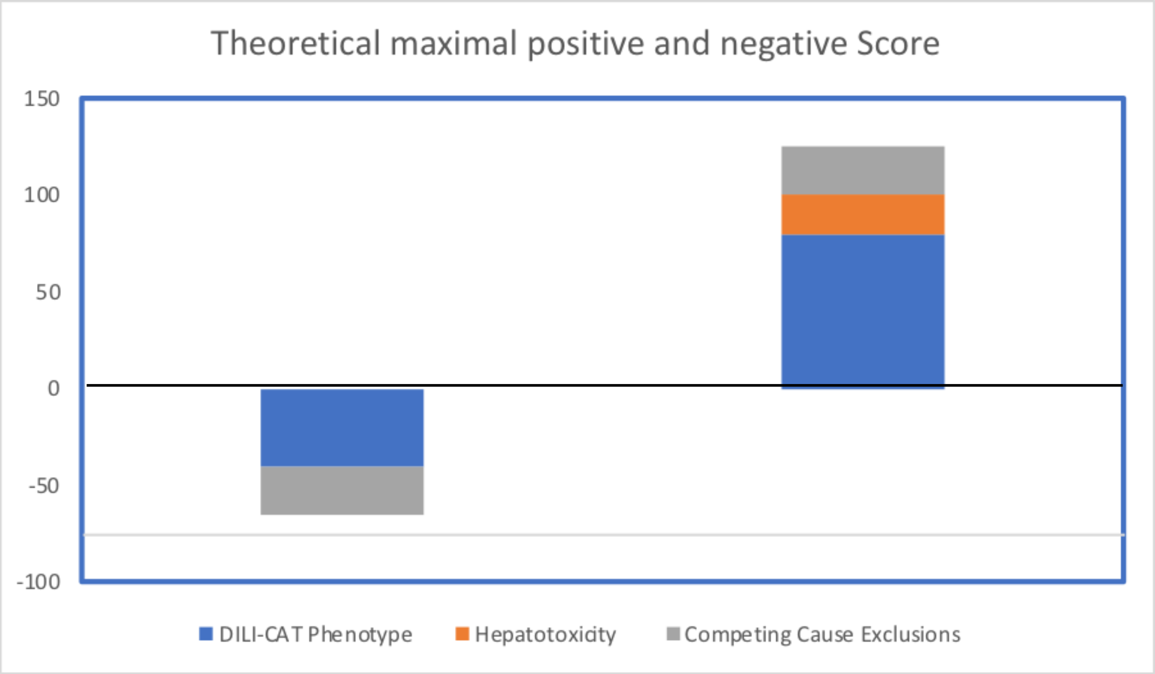


The sum score would be graded for likelihood where 95 points or higher would be "definite DILI", 75 to 95 points would be "highly likely DILI," 50 to 75 points would be "probable DILI," 25 to 50 points would be "possible DILI," and less than 25 points would be "unlikely DILI."

**Supplemental Table 1: Examples for point allocation for latency throughout the first 100 days**

| Latency (days) | Points | | | |
| --- | --- | --- | --- | --- |
| | Cyproterone | Cefazolin | AMX-CLA | PM |
| 1 | **-10** | **-10** | **-10** | 0 |
| 2 | **-10** | **-10** | -5 | 5 |
| 3 | **-10** | **-5** | -5 | 5 |
| 4 | **-10** | **-5** | 0 | 10 |
| 5 | **-10** | **-5** | 0 | 10 |
| 6 | **-10** | **-5** | 0 | 10 |
| 7 | **-10** | **-5** | 0 | 20 |
| 8 | **-10** | **-5** | 5 | 20 |
| 9 | **-10** | 10 | 10 | 20 |
| 10 | **-10** | 10 | 10 | 20 |
| 11 | **-10** | 10 | 10 | 20 |
| 12 | **-10** | 10 | 10 | 20 |
| 13 | **-10** | 10 | 10 | 20 |
| 14 | **-10** | 10 | 10 | 20 |
| 15 | **-10** | 20 | 10 | 20 |
| 16 | **-10** | 20 | 10 | 20 |
| 17 | **-10** | 20 | 20 | 20 |
| 18 | **-10** | 20 | 20 | 20 |
| 19 | -5 | 20 | 20 | 20 |
| 20 | -5 | 20 | 20 | 20 |
| 21 | -5 | 20 | 20 | 20 |
| 22 | -5 | 20 | 20 | 20 |
| 23 | -5 | 20 | 20 | 20 |
| 24 | -5 | 20 | 20 | 20 |
| 25 | -5 | 10 | 20 | 20 |
| 26 | -5 | 10 | 20 | 20 |
| 27 | -5 | 10 | 20 | 20 |
| 28 | -5 | 10 | 20 | 20 |
| 29 | -5 | 5 | 20 | 20 |
| 30 | -5 | 0 | 20 | 20 |
| 31 | -5 | -5 | 20 | 20 |
| 32 | -5 | -5 | 20 | 20 |
| 33 | 0 | -5 | 20 | 20 |
| 34 | 0 | -5 | 20 | 20 |
| 35 | 0 | -5 | 20 | 20 |
| 36 | 0 | -5 | 20 | 20 |
| 37 | 0 | -5 | 20 | 20 |
| 38 | 0 | -10 | 20 | 20 |
| 39 | 0 | -10 | 10 | 20 |
| 40 | 0 | -10 | 10 | 20 |
| 41 | 0 | -10 | 10 | 20 |
| 42 | 0 | -10 | 10 | 20 |
| 43 | 0 | -10 | 10 | 20 |
| 44 | 0 | -10 | 10 | 20 |
| 45 | 0 | -10 | 5 | 20 |
| 46 | 0 | -10 | 5 | 10 |
| 47 | 0 | -10 | 5 | 10 |
| 48 | 0 | -10 | 5 | 10 |
| 49 | 0 | -10 | 5 | 10 |
| 50 | 0 | -10 | 5 | 10 |
| 51 | 0 | -10 | 0 | 10 |
| 52 | 0 | -10 | 0 | 10 |
| 53 | 0 | -10 | 0 | 10 |
| 54 | 0 | -10 | 0 | 10 |
| 55 | 0 | -10 | 0 | 5 |
| 56 | 0 | -10 | 0 | 5 |
| 57 | 0 | -10 | 0 | 5 |
| 58 | 0 | -10 | 0 | 5 |
| 59 | 0 | -10 | 0 | 5 |
| 60 | 0 | -10 | 0 | 5 |
| 61 | 0 | -10 | 0 | 5 |
| 62 | 0 | -10 | 0 | 5 |
| 63 | 5 | -10 | 0 | 5 |
| 64 | 5 | -10 | -5 | 5 |
| 65 | 5 | -10 | -5 | 5 |
| 66 | 5 | -10 | -5 | 5 |
| 67 | 5 | -10 | -5 | 5 |
| 68 | 5 | -10 | -5 | 5 |
| 69 | 5 | -10 | -5 | 5 |
| 70 | 5 | -10 | -10 | 5 |
| 71 | 5 | -10 | -10 | 5 |
| 72 | 5 | -10 | -10 | 5 |
| 73 | 5 | -10 | -10 | 0 |
| 74 | 5 | -10 | -10 | 0 |
| 75 | 5 | -10 | -10 | 0 |
| 76 | 5 | -10 | -10 | 0 |
| 77 | 5 | -10 | -10 | 0 |
| 78 | 10 | -10 | -10 | 0 |
| 79 | 10 | -10 | -10 | 0 |
| 80 | 10 | -10 | -10 | 0 |
| 81 | 10 | -10 | -10 | 0 |
| 82 | 10 | -10 | -10 | 0 |
| 83 | 10 | -10 | -10 | 0 |
| 84 | 10 | -10 | -10 | 0 |
| 85 | 10 | -10 | -10 | 0 |
| 86 | 10 | -10 | -10 | 0 |
| 87 | 10 | -10 | -10 | 0 |
| 88 | 10 | -10 | -10 | 0 |
| 89 | 10 | -10 | -10 | 0 |
| 90 | 10 | -10 | -10 | 0 |
| 91 | 10 | -10 | -10 | 0 |
| 92 | 10 | -10 | -10 | 0 |
| 93 | 10 | -10 | -10 | 0 |
| 94 | 10 | -10 | -10 | 0 |
| 95 | 10 | -10 | -10 | 0 |
| 96 | 10 | -10 | -10 | 0 |
| 97 | 10 | -10 | -10 | 0 |
| 98 | 10 | -10 | -10 | 0 |
| 99 | 10 | -10 | -10 | 0 |
| 100 | 10 | -10 | -10 | 0 |

Bold numbers indicate respective outliers, gray background indicates numbers outside of range.
AMX/CLA, amoxicillin/clavulanate; PM, polygonum multiflorum.

### Supplemental Table 2: Relative importance of competing causes

| | **Relative importance** |
|---|---|
| **Competing causes for a hepatocellular injury pattern** | |
| a.) Hepatitis viruses A to E | |
| HAV (IgM) | 1 |
| HBV (HBcIgM) | 1 |
| HCV (anti-HCV or HCV-RNA positive) | 1 |
| HDV (IgM) (only required if HBsAg or anti-HBcIgM is positive) | 1 |
| HEV (IgM)** | 0.2 |
| b.) Alcohol abuse (AST ULN>ALT ULN × 1.5) and max AST <500 | 0.8 |
| c.) Herpesvirus: EBV, CMV or HSV or VZV | 0.3 |
| d.) Autoimmune hepatitis based on AIH Score ≥ 6* | 0.8 |
| e.) Ischemic hepatitis (evidence for hypotension) | 0.8 |
| f.) Imaging – US, CT, or MRI – biliary dilation finding only | 0.7 |
| g.) Gallstones with clinic of passage (hx of acute pain) | 0.8 |
| **Competing causes for a cholestatic injury pattern** | |
| a.) AMA (anti-mitochondrial antibodies) | 0.8 |
| b.) IgG4 levels | 0.5 |
| c.) Imaging – US, CT, MRCP, or ERCP | 1 |

* Based on ANA/SMA 1:40 or 1:80, IgG >1.0 or >1.1, histology compatible, exclusion of viral hepatitis.
** HEV IgM may deserve a higher relative value in Europe versus the United States, where the incidence of acute hepatitis E seems rarer.
AIH, autoimmune hepatitis; ALT, alanine aminotransferase; AST, aspartate aminotransferase; CMV, cytomegalovirus; CT, computerized tomography; EBV, Epstein-Barr virus; ERCP, endoscopic retrograde cholangiopancreatography; HSV, herpes simplex virus; Ig, immunoglobulin; MRCP, magnetic resonance cholangiopancreatography; ULN, upper limit of normal; US, ultrasonography; VZV, varicella zoster virus.

**Supplemental Figure 1a. Frequency distribution of R-value of cases for 4 different drugs for cyproterone (A), AMX/CLA (B), cefazolin (C), and polygonum multiflorum (D) Frequency of cases is given on the Y axis and R-value as numbers in the X axis; the vertical lines in figures A-D represent the interquartile range or 25th and 75th percentile.**

AMX/CLA, amoxicillin/clavulanate.

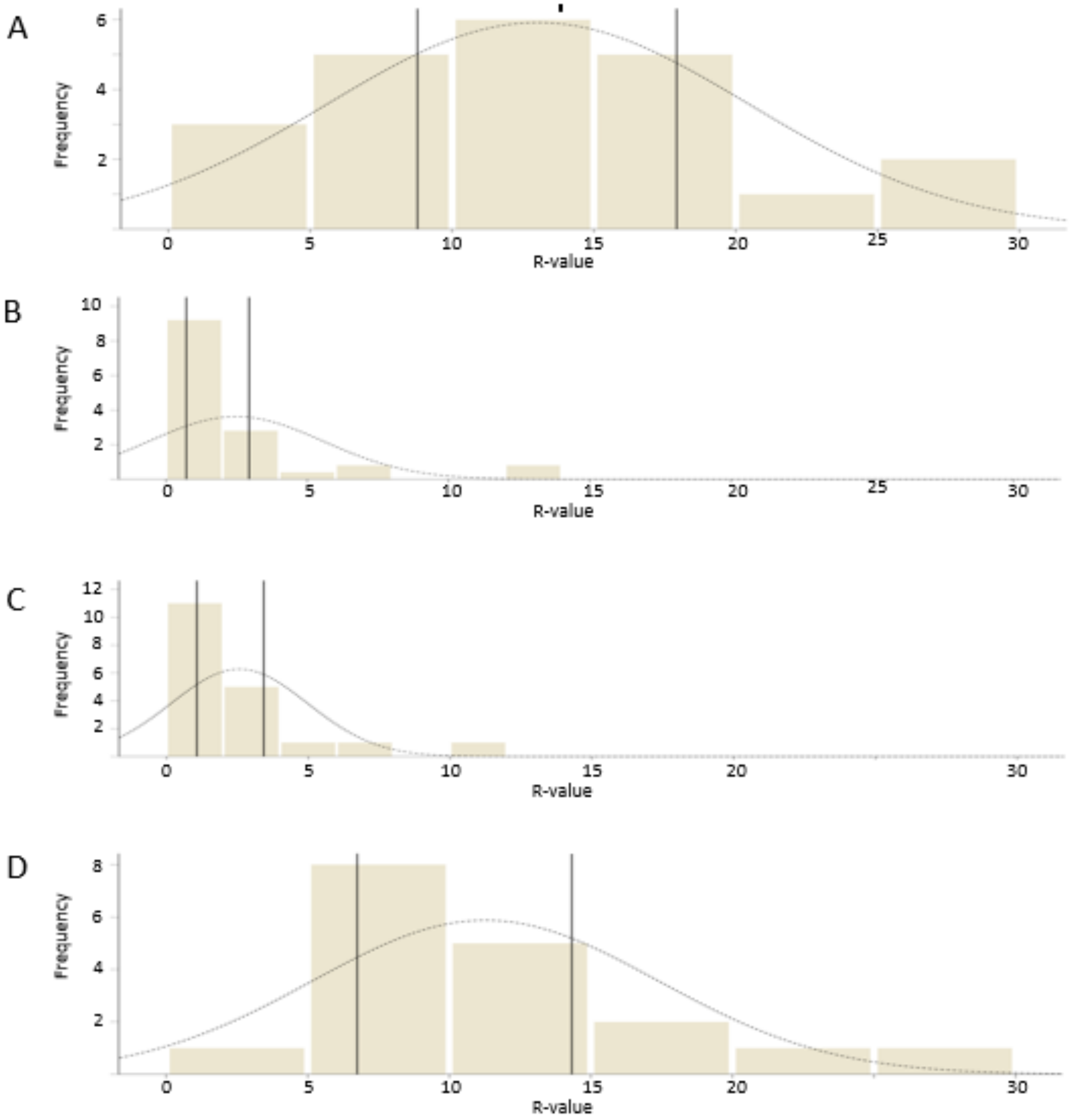

**Supplemental Figure 1b**. Frequency distribution of AST:ALT ratio of cases for 4 different drugs for cyproterone (A), AMX/CLA (B), cefazolin (C), and polygonum multiflorum (D). Frequency of cases is given on the Y axis and AST:ALT ratio as numbers in the X axis; the vertical lines in figures A-D represent the interquartile range or 25th and 75th percentile.

ALT, alanine aminotransferase; AMX/CLA, amoxicillin/clavulanate; AST, aspartate aminotransferase.

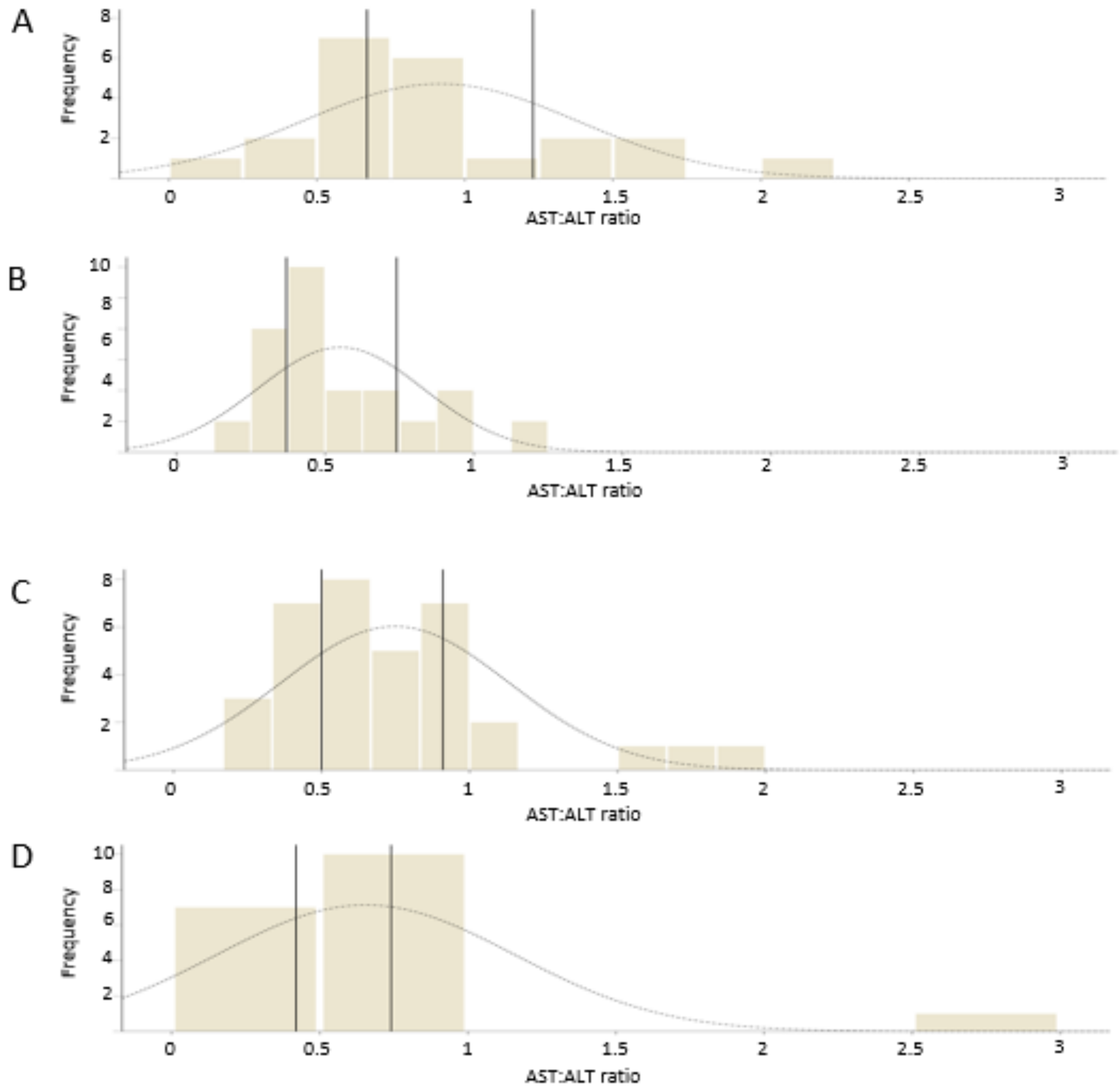

**Supplemental Figure 2a. Relative frequency of cases with various DILI-CAT scores based on cyproterone for cyproterone versus AMX/CLA, cefazolin, and polygonum multiflorum.**

AMX/CLA, amoxicillin/clavulanate; DILI-CAT, drug-induced liver injury causality assessment tool; DILI-CAT-S, drug-induced liver injury causality assessment tool score.

DILI-CAT-S$_{cytproterone}$ to assess cyproterone cases against 3 drugs

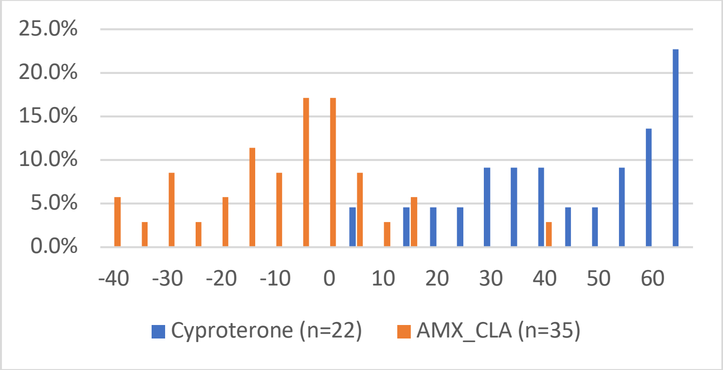


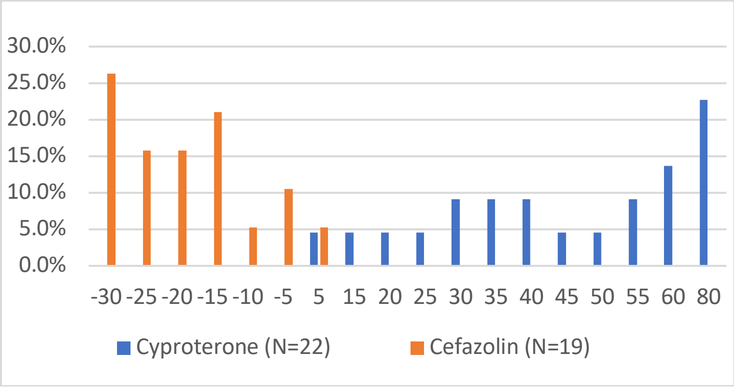


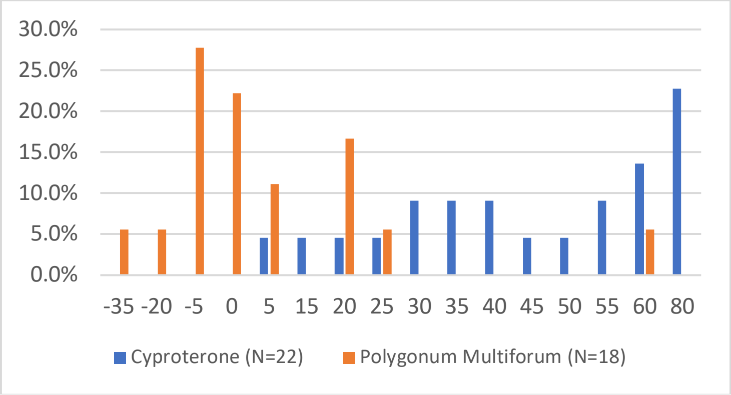

**Supplemental Figure 2b. Relative frequency of cases with various DILI-CAT scores based on AMX/CLA for AMX/CLA versus cyproterone, cefazolin, and polygonum multiflorum.**

AMX/CLA, amoxicillin/clavulanate; DILI-CAT, drug-induced liver injury causality assessment tool; DILI-CAT-S, drug-induced liver injury causality assessment tool score.

DILI-CAT-S$_{AMX-CLA}$ to assess AMX-CLA cases against 3 drugs

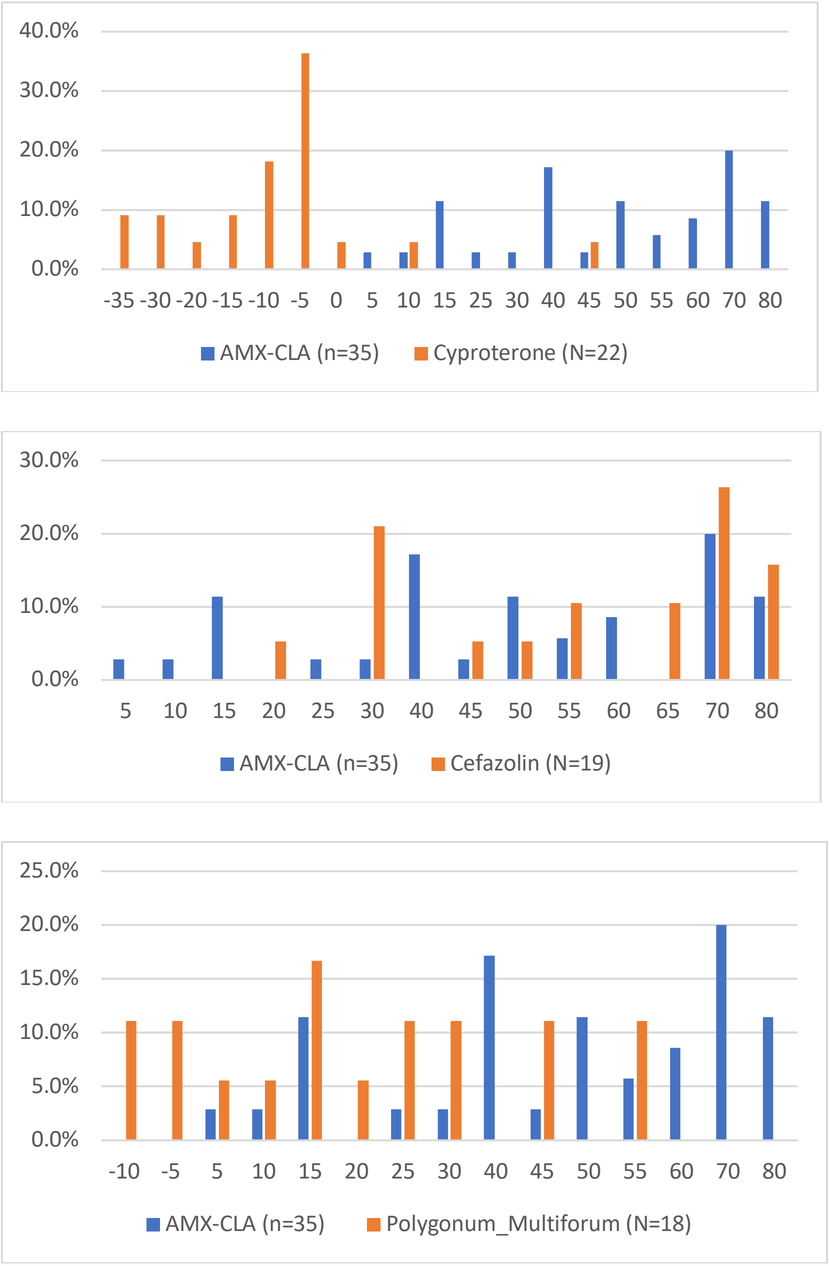

**Supplemental Figure 2c. Relative frequency of cases with various DILI-CAT scores based on cefazolin for cefazolin versus cyproterone, AMX/CLA, and polygonum multiflorum.**

AMX/CLA, amoxicillin/clavulanate; DILI-CAT, drug-induced liver injury causality assessment tool; DILI-CAT-S, drug-induced liver injury causality assessment tool score.

DILI-CAT-S$_{\text{cefazoline}}$ to assess cefazoline cases against 3 drugs

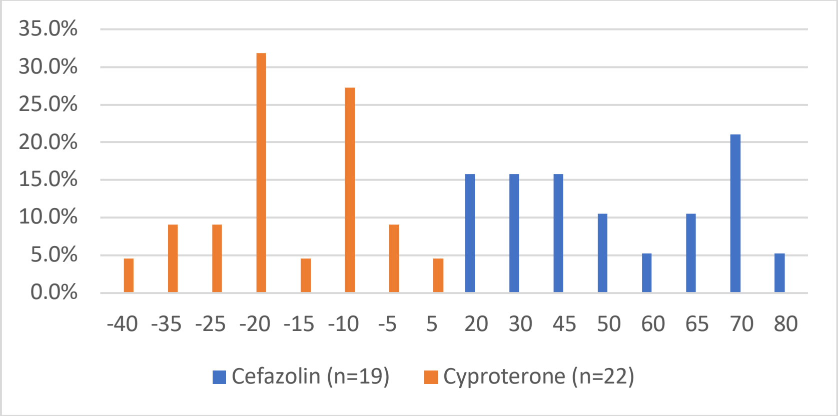


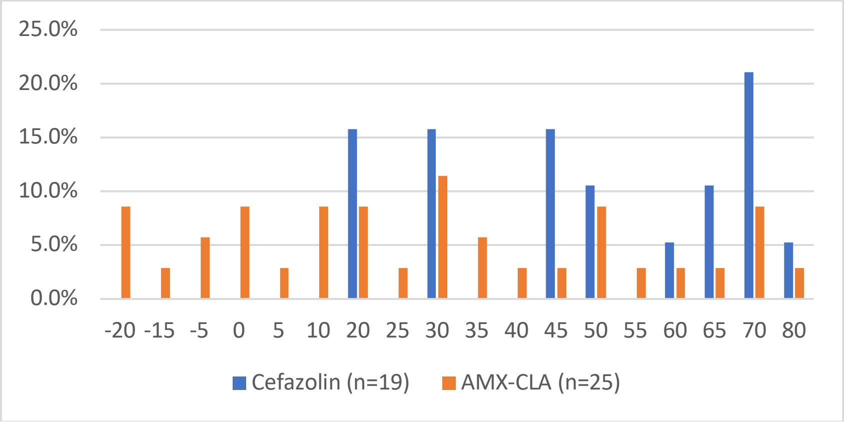


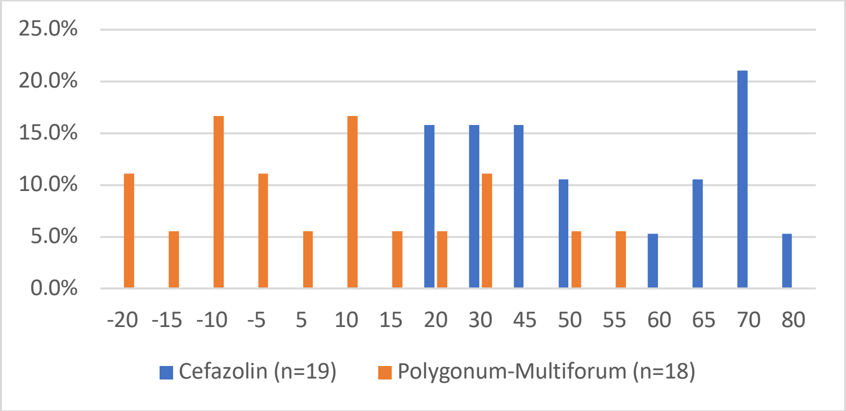

**Supplemental Figure 2d. Relative frequency of cases with various DILI-CAT scores based on polygonum multiflorum for polygonum multiflorum versus cyproterone, AMX/CLA, and cefazoline.**

AMX/CLA, amoxicillin/clavulanate; DILI-CAT, drug-induced liver injury causality assessment tool; DILI-CAT-S, drug-induced liver injury causality assessment tool score.

DILI-CAT-S$_{polygonum\text{-}multiforum}$ to assess polygonum multiflorum cases against 3 drugs

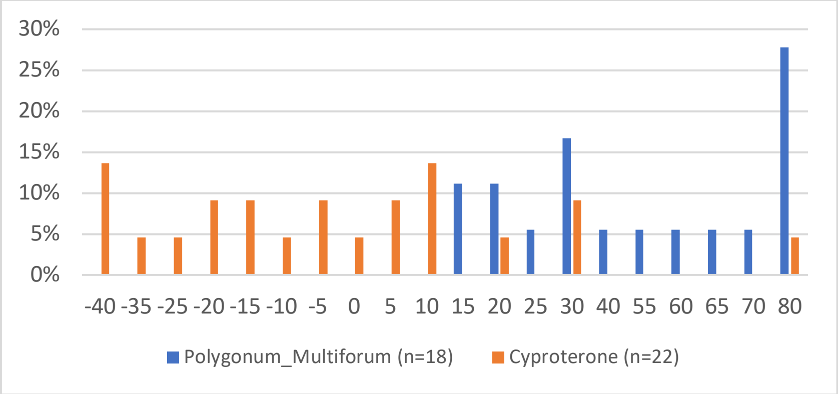


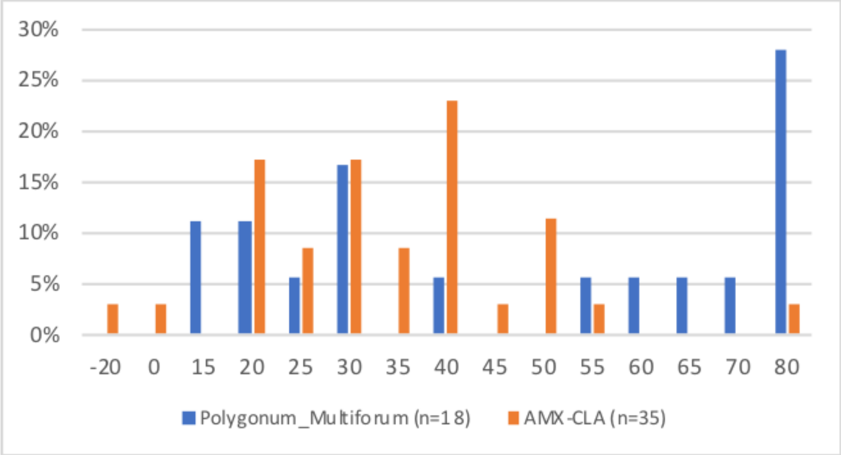


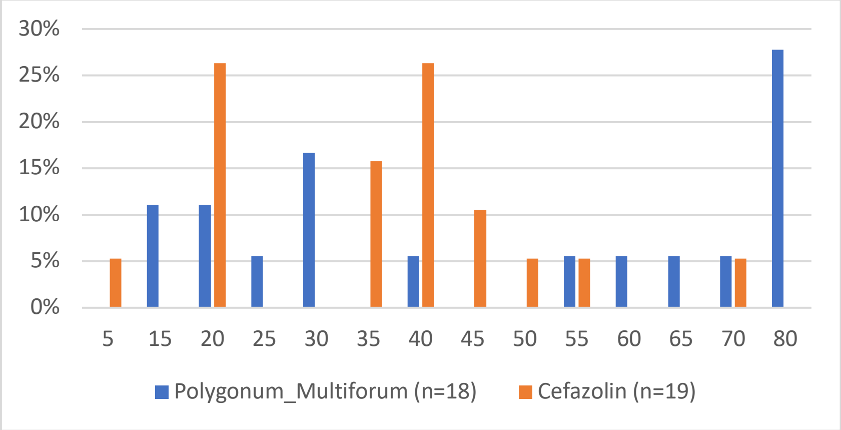
